# Evaluation of gait cycle time variability in patients with knee osteoarthritis using a triaxial accelerometer

**DOI:** 10.1101/2021.07.02.21259710

**Authors:** Takeshi Akimoto, Kenji Kawamura, Takaaki Wada, Naomichi Ishihara, Akane Yokota, Takehiko Suginoshita, Shigeki Yokoyama

**Affiliations:** Graduate School of Health Science, Kibi International University, Takahashi-shi, Okayama, Japan; Department of Orthopedics, Medical Corporation Suginoshita Orthopedic Clinic, Rokujizo, Uji-shi, Kyoto, Japan; Graduate School of Health Science, Kyoto Tachibana University, Yamashina-ku, Kyoto-shi, Kyoto, Japan

## Abstract

Knee osteoarthritis can alter gait variability. However, few studies have compared the temporal factors of the gait cycle between patients with knee osteoarthritis and healthy subjects. Furthermore, no studies have investigated the relationship between gait variability and potential contributing factors (knee joint functions such as muscle strength) in knee osteoarthritis. The first objective of this study was to compare gait cycle variability between female patients with knee osteoarthritis and healthy elderly women to determine gait characteristics in patients with knee osteoarthritis. The second objective was to examine whether gait cycle variability in knee osteoarthritis is associated with potential contributing factors. Twenty-four female patients diagnosed with knee osteoarthritis and 12 healthy elderly women participated. Gait cycle variability (coefficient of variation of gait cycle time), knee extension range of motion, knee extension strength, 5-meter walk test, Timed Up & Go Test, and Western Ontario and McMaster Universities Osteoarthritis Index were measured. All assessment results were compared between the knee osteoarthritis and healthy groups. Gait cycle variability was significantly higher in the knee osteoarthritis group (3.2%±1.5%) compared to the healthy group (2.1%±0.7%). A significant positive correlation was found between the gait cycle variability and 5-meter walk test (r=0.46) and Western Ontario and McMaster Universities Osteoarthritis Index (r=0.43). The gait of patients with knee osteoarthritis may be more unstable than that of healthy individuals. In addition, unstable gait may be associated with gait speed and quality of life. Therefore, we believe that rehabilitation to improve unstable gait can enhance the quality of life of patients with knee osteoarthritis.

## Introduction

Knee osteoarthritis (OA) is a common disease that imposes an enormous personal and social burden. In Japan, Yoshimura et al. [1] reported that the prevalence of radiographic knee OA was 42.6% in men and 62.4% in women aged >40 years, indicating that knee OA is an epidemiologically important disease. Knee OA is a common disease in elderly women [2] and is a leading cause of pain and dysfunction [3]. The main symptom is decreased gait ability, which can have a negative impact on activities of daily living and quality of life [4]. Therefore, it is important to accurately assess the gait ability of patients with knee OA and improve it through rehabilitation.

It has been widely reported that gait function is diminished by knee OA. Spatiotemporal gait parameters such as slower gait speed, shorter stride length, increased stride time, increased stance phase duration, and increased double support time are worsened [5–8]. In addition to these parameters, knee OA has recently been shown to alter gait variability.

Research on gait variability in patients with knee OA is ongoing. Gait variability has been compared between patients with knee OA and healthy subjects and between different Kellgren–Lawrence severity levels, and outcomes have differed in each study [9–13]. In particular, studies focusing on the variability of spatiotemporal parameters have investigated the standard deviation and coefficient of variation (CV) of gait cycle time and stance time.

Clermont and Tanimoto et al. [9, 10] reported that there was no significant difference in the gait cycle time standard deviation or CV between the knee OA and healthy groups. However, Kiss et al. [11] reported that the knee OA group had a significantly higher CV of stance time. Oka et al. [12] reported that there was a significant difference in the gait cycle time CV between knee OA patients with a fear of falling and those without a fear of falling. Thus, studies focusing on the gait variability of spatiotemporal parameters in knee OA have not yielded a consistent view.

On the other hand, many researchers have investigated gait variability in healthy elderly people. Hausdorff and Balasubramanian et al. [14, 15] reported that gait variability is related to gait speed in community-dwelling older adults. In addition, Bogen et al. [16] reported that gait variability tended to be related to muscle strength measured two years earlier. Matsuda et al. [17] suggested that muscle strength must be improved to reduce gait variability. Thus, studies have investigated the relationship between gait variability and potential contributing factors in elderly people in the community. However, to the best of our knowledge, no study has investigated the relationship between gait variability and potential contributing factors (knee joint functions such as muscle strength, range of motion, and physical functions such as gait speed) in knee OA.

The first objective of this study was to compare gait cycle variability between female patients with knee OA and healthy elderly women to determine gait characteristics in patients with knee OA. The second objective was to examine whether gait cycle variability in knee OA is associated with potential contributing factors. We hypothesized that 1) gait cycle variability would be different in knee OA and healthy participants and 2) potential contributing factors such as muscle strength would be associated with gait cycle variability.

## Materials and Methods

### Participants

This study recruited participants in two groups: patients with knee OA and healthy elderly women. Twenty-four female patients diagnosed with knee OA by radiography were included in the knee OA group [age: 70.8±5.7 years, height: 1.56±0.05 m, weight: 56.6±6.4 kg, body mass index (BMI): 23.3±2.4 kg/m^2^], and 12 healthy elderly women living in the community were included in the healthy group (age: 69.8±8.1 years, height: 1.53±0.06 m, weight: 53.5±6.1 kg, BMI: 22.9±2.9 kg/m^2^). The Kibi International University Ethics Committee approved all measures of this study (approval number: 19-14). All participants provided written informed consent before participating in the study.

The criteria for inclusion in the knee OA group were (1) women and (2) patients who were able to participate in rehabilitation at least once a week. The exclusion criteria were (1) severe pain other than knee pain, (2) history of lower extremity trauma or surgery, (3) history of serious cardiac or pulmonary disease, and (4) history of rheumatoid arthritis. The inclusion criteria for the healthy group were as follows: (1) women. The exclusion criteria were (1) pain in the lower limbs, (2) a history of lower extremity trauma or surgery, (3) a history of serious cardiac or pulmonary disease, and (4) a history of rheumatoid arthritis.

The Kellgren–Lawrence classification of the knee OA group was grade I (No patient), grade II (15 patients), grade III (8 patients), and grade IV (1 patient).

### Measurement Method

Prior to the trial task, participants were given practice time to become accustomed to gait on the treadmill. The comfortable gait speed for each participant was determined during the practice period. This comfortable gait speed was used for data collection in this study. After an adequate rest period, the participants walked on the treadmill for 1 minute at a comfortable gait speed (comfortable speed was 2.3±0.8 km/h for the knee OA group and 2.5±0.8 km/h for the healthy group). During the task, the rating of the perceived exertion scale (Borg’s 6–20 scale) was assessed to investigate exercise intensity [18].

A triaxial accelerometer (TSND121, ATR-Promotions Co., Kyoto, Japan) was used to collect data during the trial. The size of the sensor was 37 mm×46 mm×12 mm and weighed 22 g. A triaxial accelerometer was attached to the third lumbar vertebra of each participant. The acceleration waveform data during gait were transmitted to the computer via Bluetooth. The raw sensor data were sampled at a frequency of 100 Hz. The heel ground contact of the gait was analyzed using acceleration waveform information. The duration of one gait cycle was defined as the time between heel ground contact and the next heel ground contact on the same side. The CV, which is the ratio of the standard deviation to the mean, was calculated from the obtained gait cycle time. This CV was defined as the gait cycle variability and was used in the results.

To evaluate knee joint function, knee extension range of motion (ROM) and knee extension strength were measured. Knee extension ROM was measured in the supine position using a goniometer. Knee extension strength was measured as the isometric strength at 90° of knee flexion. A hand-held dynamometer (μTas F-1, Anima Co., Tokyo, Japan) was used to measure muscle strength.

The 5-m walk test (5MWT) [19] and the Timed Up & Go Test (TUG) [20] were used to assess gait ability. The 5MWT measured the gait speed over a distance of 5 meters. An 11-meter gait path was used, with 3 meters at each end prepared for acceleration and deceleration and the central 5 meters used for measurement. The participants were instructed to walk as quickly as possible. TUG measured the time it took for a participant to get up from a chair, walk 3 meters, turn around, walk back to the chair, and sit down.

The Western Ontario and McMaster Universities Osteoarthritis Index (WOMAC) [21] was used to assess pain, stiffness, and function. The WOMAC was used as a specific quality of life measure for knee OA. Higher WOMAC scores indicate more severe functional limitations.

### Statistical Analysis

The statistical software IBM SPSS for Windows version 26 was used for the statistical analysis. Physical characteristics, gait cycle variability, gait speed on the treadmill, Borg’s 6–20 scale, knee extension strength, knee extension ROM, 5MWT, TUG, and WOMAC were compared between the knee OA and healthy groups. Normality was checked using the Shapiro-Wilk test, and either the t-test or Mann-Whitney U test was performed. The effect size r was calculated from the T-value (for t-test) and Z-value (for Mann-Whitney U test) using Microsoft Excel.

In addition, the relationships between gait cycle variability and knee extension strength, knee extension ROM, 5MWT, TUG, and WOMAC in the knee OA group were examined. Normality was checked using the Shapiro-Wilk test, and Spearman’s rank correlation coefficient was used. Correlation results were interpreted as negligible (p<0.30), weak (p=0.30–0.50), moderate (p=0.50–0.70), high (p=0.70–0.90), or very high (p>0.90).

The sample size was determined after conducting a pilot study with 12 participants (6 in the OA group and 6 in the healthy group). The allocation ratio was 2:1 for the knee OA and healthy groups, and the significance (α) and power were set at 0.05 and 0.8, respectively. The calculated sample size was 20 for the OA group and 10 for the healthy group, and we were able to recruit a sufficient number of participants.

## Results

The participants’ characteristics for the knee OA and healthy groups are shown in Table 1. There were no significant differences in age, height, weight, or BMI between the knee OA and healthy groups.

**Table 1.** Participant characteristics.

|  | Knee OA group<br>(n=24) | Healthy group (n=12) | p-value | Effect size r |
| --- | --- | --- | --- | --- |
| Age (years) | 70.8 (5.7) | 69.8 (8.1) | p=0.80 | 0.05 |
| Height (m) | 1.56 (0.05) | 1.53 (0.06) | p=0.14 | 0.25 |
| Body mass (kg) | 56.6 (6.4) | 53.5 (6.1) | p=0.38 | 0.15 |
| BMI (kg/m<sup>2</sup>) | 23.3 (2.4) | 22.9 (2.9) | p=0.68 | 0.07 |
| KL Score (n) |  |  |  |  |
| Grade 0 | 0 |  |  |  |
| Grade 1 | 15 |  |  |  |
| Grade 2 | 8 |  |  |  |
| Grade 3 | 1 |  |  |  |

Data are presented as mean ( ± SD). OA: osteoarthritis; BMI: body mass index; KL:
Kellgren–Lawrence

Gait cycle variability was 3.2±1.5% in the knee OA group and 2.1±0.7% in the healthy group, which was significantly higher in the knee OA group (medium effect size, 0.44). The comfortable gait speed on the treadmill was 2.3±0.8 km/h in the knee OA group and 2.5±0.8 km/h in the healthy group, which was not significantly different. The Borg’s 6– 20 scale during the gait task was 11.0±2.3 in the knee OA group and 11.1±1.3 in the healthy group and both groups fell into the “fairly light” category, with no significant difference (Table 2).

**Table 2.** Comparison of gait on the treadmill, knee function, gait ability, and WOMAC between the knee OA group and healthy group.

|  | Knee OA<br>group<br>(n=24) | Healthy group<br>(n=12) | p-value | Effect size<br>r |
| --- | --- | --- | --- | --- |
| **Gait on the treadmill** |  |  |  |  |
| Gait cycle variability (%) | 3.2 (1.5) | 2.1 (0.7) | p<0.01 | 0.44 |
| Gait speed on the treadmill<br>(km/h) | 2.3 (0.8) | 2.5 (0.8) | p=0.56 | 0.10 |
| Borg's 6-20 scale | 11.0 (2.3) | 11.1 (1.3) | p=0.96 | 0.01 |
| **Knee function** |  |  |  |  |
| Knee extension ROM (°) | -4.2 (4.3) | -0.8 (1.9) | p<0.05 | 0.41 |
| Knee extension strength (kgf/kg) | 0.38<br>(0.09) | 0.51 (0.12) | p<0.00<br>1 | 0.52 |
| **Gait ability** |  |  |  |  |
| 5MWT (s) | 3.8 (0.9) | 2.9 (0.3) | p<0.00<br>1 | 0.55 |
| TUG (s) | 6.8 (1.0) | 5.9 (0.4) | p<0.01 | 0.43 |
| **Assessments of pain, stiffness, and function** |  |  |  |  |
| WOMAC | 20.5 (9.0) | 4.3 (7.4) | p<0.00<br>1 | 0.69 |

Data are presented as mean ( ± SD). WOMAC: Western Ontario and McMaster Universities Osteoarthritis Index; OA: osteoarthritis; ROM: range of motion; 5MWT: 5-meter walk test; TUG: Timed Up & Go test;

The results of the other measurements are presented in Table 2.

Knee function was significantly lower in the knee OA group in terms of both extensor muscle strength and extension ROM (medium to large effect size, 0.41–0.52). The 5MWT and TUG test results were significantly slower in the knee OA group (medium to large effect size, 0.43–0.55). WOMAC scores were significantly higher in the knee OA group (large effect size, 0.69). Furthermore, Table 3 shows the results of the correlations between gait cycle variability and knee extension strength, knee extension ROM, 5MWT, TUG, and WOMAC in the knee OA group. A significant weak positive correlation was found between gait cycle variability and 5MWT (r=0.46). In addition, there was a significant weak positive correlation between gait cycle variability and WOMAC scores (r=0.43). There was no significant association between gait cycle variability and other factors (Table S1).

**Table 3.** Potential contributing factors associated with gait cycle variability in the knee OA group.

|  | Knee OA group (n=24) |  |
| --- | --- | --- |
| | Spearman's rank correlation coefficient ($\rho$) | p-value |
| Knee extension ROM | −0.25 | p=0.23 |
| Knee extension strength | −0.24 | p=0.25 |
| 5MWT | 0.46 | p<0.05 |
| TUG | 0.33 | p=0.11 |
| WOMAC | 0.43 | p<0.05 |

OA: osteoarthritis; ROM: range of motion; 5MWT: 5-meter walk test; TUG: Timed Up & Go test; WOMAC: Western Ontario and McMaster Universities Osteoarthritis Index

## Discussion

The first objective of this study was to investigate whether gait cycle variability differed between the knee OA and healthy groups. The results showed that the gait cycle variability of the knee OA group was significantly larger than that of the healthy group. This study showed that gait cycle variability may play an important role in the rehabilitation of patients with knee OA.

There were no significant differences in age, height, weight, or BMI between the knee OA and healthy groups (Table 1). In other words, there was no difference in the physical characteristics between the two groups, and only the knee OA group had OA symptoms such as knee pain. Therefore, the participants in this study were suited to the purpose of the study, which was to compare patients with knee OA to healthy individuals.

Gait cycle variability was significantly greater in the knee OA group than in the healthy group. Kiss et al. [11] reported a significantly greater stance time CV in the knee OA group than in the healthy group. Therefore, among the studies comparing the CV of spatiotemporal gait parameters, the present study supports the work of Kiss et al. [11]. Kiss et al. [11] used the same gait speed for all participants when collecting data on treadmill gait. On the other hand, the present study collected data at a comfortable gait speed for each participant. A new finding was that the gait variability of spatiotemporal parameters in the knee OA group was greater than that in the healthy group, even at the participants’ daily gait speed. Furthermore, previous studies have reported that increased gait variability is associated with an increased risk of falls [14, 22, 23]. Although the present study did not investigate falls, the gait of patients with knee OA may be less stable than that of healthy individuals.

Borg’s 6–20 scale for treadmill gait showed no significant difference between the two groups (Table 2). Therefore, in terms of exercise intensity, the gait task speed was appropriate because participants in both groups fell into the “fairly light” category and performed the task at the same intensity.

Knee joint function, gait speed, and WOMAC scores were lower in the knee OA group than in the healthy group (Table 2). Previous studies have also reported reduced knee extensor strength and gait speed in patients with knee OA compared to those in healthy subjects [6–8, 24, 25], which means that the results of the present study support those of previous studies. Furthermore, there were significant weak positive correlations between gait cycle variability and 5MWT (r=0.46) and between gait cycle variability and WOMAC (r=0.43). Gait cycle variability has been reported to correlate with gait speed in healthy participants [14, 15]. The present study showed that gait cycle variability was associated with gait speed, even in patients with knee OA. The correlation of gait cycle variability with WOMAC indicates that gait variability reflects the quality of life and physical function of patients with knee OA. Kalsi-Ryan et al. [26] reported a correlation between Japan Orthopedic Association score and gait CV in patients with spondylolisthesis osteoarthritis. Correlation with such disease-specific assessments of physical functioning, even in patients with knee OA, indicates that gait variability is associated with quality of life. Therefore, reducing gait CV may lead to improved quality of life.

On the other hand, no relationship was found between knee function and gait cycle variability. This result differs from our hypothesis. It has been reported that there is an association between gait cycle variability and knee extension strength in healthy elderly people [14, 16, 17]. There was a difference in the results for knee OA. To the best of our knowledge, no previous study has examined the relationship between gait cycle variability and extensor muscle strength in knee OA. Therefore, different factors may be associated with gait cycle variability in patients with knee OA than in healthy older adults. In a previous study of local elderly people, a correlation between hip abduction strength and gait cycle variability was reported [27]. In this study, only knee extension strength was evaluated; therefore, it is necessary to evaluate knee flexion strength and hip joint strength in the future.

## Research limitations

This study has some limitations. First, the study included 24 participants in the knee OA group and 12 in the healthy group, which is a small sample size. The sample size may have affected the results. Second, the trial task was gait on a treadmill. Although the participants were given time to practice, it is possible that their experience with the treadmill may have influenced the results. One of the reasons for adopting the treadmill gait was to increase the number of steps. Lord et al. [28] reported that it is possible to measure gait cycle variability even at 10 m walking, but in many cases, a certain number of steps is ensured, such as measuring 100 gait cycles or 6–10 minutes of walking [29, 30]. Therefore, we adopted the treadmill gait to increase the number of steps. However, treadmill gait has a disadvantage in that it is different from a normal gait. In the future, it will be necessary to consider the design of the study to increase the number of steps with a continuous gait in a large space.

## Conclusion

In our study, we compared differences in gait variability between female patients with knee OA and healthy elderly women. The gait of patients with knee OA may be more unstable than that of healthy individuals. In addition, unstable gait may be associated with gait speed and quality of life. Therefore, we believe that rehabilitation to improve unstable gait can enhance the quality of life of patients with knee OA.

## Data Availability

All relevant data are within the manuscript and its Supporting information files.

## Acknowledgments

We would like to thank Editage (www.editage.com) for English language editing.

## Supporting information

**S1 Table. Research Data**

